# SARS-CoV-2 infection does not significantly cause acute renal injury: an analysis of 116 hospitalized patients with COVID-19 in a single hospital, Wuhan, China

**DOI:** 10.1101/2020.02.19.20025288

**Authors:** Luwen Wang, Xun Li, Hui Chen, Shaonan Yan, Yan Li, Dong Li, Zuojiong Gong

## Abstract

**Background:** Whether the patients with COVID-19 infected by SARS-CoV-2 would commonly develop acute renal function damage is a problem worthy of clinical attention. This study aimed to explore the effects of SARS-CoV-2 infection on renal function through analyzing the clinical data of 116 hospitalized COVID-19-confirmed patients.

**Methods:** 116 hospitalized COVID-19-confirmed patients enrolled in this study were hospitalized in the Department of Infectious Diseases, Renmin Hospital of Wuhan University from January 14 to February 13, 2020. The recorded information includes demographic data, medical history, contact history, potential comorbidities, symptoms, signs, laboratory test results, chest computer tomography (CT) scans, and treatment measures. SARS-CoV-2 RNA in 53 urine sediments of enrolled patients was examined by real-time RT-PCR.

**Findings:** 12 (10.8%) and 8 (7.2%) patients showed mild elevation of blood urea nitrogen or creatinine, and trace or 1+ albuminuria respectively in 111 COVID-19-confirmed patients without basic kidney disease. In addition, 5 patients with chronic renal failure (CRF) were undergone regular continuous renal replacement therapy (CRRT) were confirmed infection of SARS-CoV-2, and diagnosed as COVID-19. Beside the treatment of COVID-19, CRRT was also applied three times weekly. The course of treatment, the renal function indicators showed stable, without exacerbation of CRF, and pulmonary inflammation was gradually absorbed. All 5 patients with CRF were survived. Moreover, SARS-CoV-2 RNA in urine sediments was positive only in 3 patients from 48 cases without renal illness before, and one patient had a positive for SARS-CoV-2 ORF 1ab from 5 cases with CRF.

**Interpretation:** Acute renal impairment was uncommon in COVID-19. SARS-CoV-2 infection does not significantly cause obvious acute renal injury, or aggravate CRF in the COVID-19 patients.

## Introduction

In December 2019, an acute respiratory infectious disease caused by a novel coronavirus occurred in Wuhan, Hubei Province, China, which is now officially named as “2019 coronavirus disease (COVID-19)” by the WHO ^[1-3]^. The disease has spread rapidly from Wuhan to other regions in China. As of 24:00 on February 13, 2020, a total of 63,851 COVID-19-confirmed cases were reported in China ^[4]^. Internationally, cases have been reported in 24 countries and 5 continents ^[5]^. On January 3, 2020, a novel coronavirus was identified in a bronchial alveolar lavage fluid sample from a patient in Wuhan and confirmed to be the cause of COVID-19 ^[6]^. Whole genome sequencing and systematic analysis showed that this novel coronavirus is a distinct clade from beta coronavirus associated with human severe acute respiratory syndrome (SARS) and Middle East respiratory syndrome (MERS) ^[6]^, which was officially named “SARS-CoV-2” by WHO now. Although the origin of SARS-CoV-2 is still being investigated, current evidence suggests it was transmitted to humans through the spread of wild animals illegally sold in Huanan Seafood Wholesale Market ^[7]^. Case reports have confirmed the interpersonal transmission of SARS-CoV-2 ^[8]^. Currently, there are no specific treatments or vaccines for COVID-19. Huang et al. first reported 41 COVID-19 cases, most of whom had a history of exposure to Huanan Seafood Wholesale Market ^[9]^. The patient’s clinical manifestations included fever, unproductive cough, dyspnea, myalgia, fatigue, normal or decreased white blood cell count, and imaging evidence of pneumonia ^[9]^. Wang et al. reported in 138 hospitalized COVID-19-confirmed cases, presumed hospital-related transmission of SARS-CoV-2 was suspected in 41% of patients, 26% of patients received ICU care, and mortality was 4.3% ^[10]^. In this study, the clinical data of 116 hospitalized COVID-19-confirmed patients were analyzed, and the effects of SARS-CoV-2 infection on renal function were explored.

## Methods

### Study design and participants

Renmin Hospital of Wuhan University is located in Wuhan City, Hubei Province, an area where COVID-19 is endemic. It is one of the city’s major tertiary teaching hospitals and the designated COVID-19-treatment hospital by the Wuhan Municipal Government. In this study, 116 COVID-19-confirmed patients were enrolled, who were hospitalized in the Department of Infectious Diseases, Renmin Hospital of Wuhan University from January 14 to February 13, 2020 (before January 20, they were diagnosed as unknown origin viral pneumonia). Oral consent was obtained from all patients. A confirmed-diagnosis of all COVID-19 patients participating in this study was made according to the WHO’s interim guidelines ^[11]^. This study was approved by the Institutional Ethics Committee of Renmin Hospital of Wuhan University.

### Nucleic acid detection of SARS-CoV-2

All COVID-19 patients enrolled in this study were laboratory-confirmed cases, which were identified with nucleic acid detection of SARS-CoV-2 from a throat swab samples using reverse transcription-polymerase chain reaction (RT-PCR). The criteria for the confirmed-diagnosis of SARS-CoV-2 was that at least one gene site was amplified to be positive for nucleocapsid protein (NP) gene and open reading frame (ORF) 1ab gene. In brief, the throat swab was put into a collection tube containing 150μl viral preservation solution, and the total RNA was extracted within 2h with the respiratory sample RNA separation Kit (Zhongzhi, Wuhan). The suspension was used for RT-PCR assay of SARS-CoV-2 RNA. Two target genes, including NP and ORF1ab, were simultaneously amplified and tested during the real-time RT-PCR assay. Target 1 (NP): forward primer GGGGAACTTCTCCTGCTAGAAT; reverse primer CAGACATTTTGCTCTC AAGCTG; and the probe 5’-FAM-TTGCTGCTGCTTGACAGATT-TAMRA-3’. Target 2 (ORF1ab): forward primer CCCTGTGGGTTTTACACTTAA; reverse primer ACGATTGTGC ATCAGCTGA; and the probe 5’-VIC-CCGTCTGCGGTATGTGGAAAGGTTATGG-BHQ1 -3’. The real-time RT-PCR assay was performed using a 2019-nCoV nucleic acid detection kit according to the manufacturer’s protocol (Shanghai bio-germ Medical Technology Co Ltd). Specific primers and probes for SARS-CoV-2 RNA detection were based on the recommendation by the National Institute for Viral Disease Control and Prevention (China) (http://ivdc.chinacdc.cn/kyjz/202001/t20200121_211337.html).

### Data collection

Epidemiological, clinical, laboratory, and radiological characteristics were recorded. The patients’ medical history as well as treatment and outcome data, were also obtained through data collection tables in electronic medical records. Data were reviewed by a team of specialists. The recorded information includes demographic data, medical history, contact history, potential comorbidities, symptoms, signs, laboratory test results, chest computer tomography (CT) scans, kidney B-ultrasonic examination and treatment measures (i.e. antiviral therapy, glucocorticoid usage, breathing support, kidney replacement therapy). The onset date was defined as the date on which symptoms appear. Different clinical categories were defined for all COVID-19 patients participating in the study according to the WHO’s interim guidelines, including Mild pneumonia, Severe pneumonia, and Acute Respiratory Distress Syndrome (ARDS) ^[11]^. Acute kidney injury was identified according to Kidney Disease: Improving Global Outcomes (KDIGO) ^[12]^.

### Statistical analysis

Categorical variables were described as frequency and percentage, and continuous variables were described as using mean, median, and interquartile range (IQR) values. When the data were normally distributed, independent *t*-tests were used to compare the mean of continuous variables. Otherwise, the Mann-Whitney test is used. Although Fisher’s exact test was used with limited data, the χ2 test was used to compare the proportion of categorical variables. All statistical analyses were performed using SPSS version 13.0 software. A *P* value of less than 0.05 is statistically significant.

## Results

### Presenting Characteristics

In this study, the median age of 116 COVID-19-confirmed patients was 54 years (IQR, 38-69; range 20-95 years), of which 67 (57.8%) were male. In these patients, 59 (50.8%) were mild pneumonia, and 46 (39.7%) were severe pneumonia, who entered the isolation ward, while 11 (9.5%) were ARDS, who were transferred to intensive care unit (ICU) (Table 1). In these patients, 51 (43.9%) cases had one or more comorbidities. Hypertension (43 [37.1%]), diabetes (18 [15.5%]), malignant tumors (12 [10.3%]), cerebral infarction (7 [6.0%]), and chronic renal failure (CRF) with long-term hemodialysis (5 [4.3%]) were the common coexisting diseases.

**Table 1.** Baseline Characteristics of 116 COVID-19-confirmed patients [*n* (%)]

|  | Total<br>(n=116) | clinical categories of pneumonia |  |  | *P* value |
| --- | --- | --- | --- | --- | --- |
|  |  | Mild<br>(n=59) | Severe<br>(n=46) | ARDS<br>(n=11) |  |
| Age, median (IQR), y | 54 (38-69) | 45 (27-56) | 52 (35-64) | 67 (58-81) | <0.001 |
| Gender |  |  |  |  |  |
| Male | 67 (57.8) | 34 (57.6) | 27 (58.7) | 6 (52.0) | 1.000 |
| Female | 49 (42.2) | 25(42.4) | 19 (41.3) | 5 (45.5) | 1.000 |
| Comorbidities |  |  |  |  |  |
| Hypertension | 43 (37.1) | 23 (38.9) | 15 (32.6) | 5 (45.5) | 0.533 |
| diabetes | 18 (15.5) | 8 (13.6) | 6 (13.0) | 4 (36.4) | 0.067 |
| malignant tumors | 12 (10.3) | 1 (1.7) | 5 (10.9) | 6 (52.0) | <0.001 |
| cerebral infarction | 7 (6.0) | 1 (1.7) | 4 (8.7) | 2 (18.2) | 0.132 |
| chronic renal failure | 5 (4.3) | 0 | 5 (10.9) | 0 | 1.000 |

*P* values indicate differences between ARDS and non-ARDS patients. *P* <0.05 was considered statistically significant.

### Changes of kidney-related clinical indicators

As shown in Table 2, 111 COVID-19-confirmed patients without kidney basic disease did not develop obvious abnormal renal function after infection with SARS-CoV-2 and during the treatment of pneumonia. Although 12 (10.8%) patients without kidney basic disease showed mild elevation of blood urea nitrogen or creatinine, and 8 (7.2%) patients without kidney basic disease showed trace or 1+ albuminuria after infection with the virus and during the treatment of pneumonia, they gradually returned to normal after a follow-up and did not receive special treatment for the kidneys. At present, none of the patients exhibited acute renal failure (ARF). In addition, the patients with chronic renal failure (CRF) were still undergoing regular continuous renal replacement therapy (CRRT) except for the treatment of COVID-19. During the course of treatment, the monitoring of renal function indicators showed stable, without exacerbation of CRF, and re-examination of CT showed that pulmonary inflammation was gradually absorbed.

**Table 2.** Changes of kidney function in 116 COVID-19-confirmed patients.

| COVID-19-confirmed patients<br>(n=116) | n | BUN (mmol/L)<br>3.6 ~ 9.5 | Cr (μmol/L)<br>57 ~ 111 | eGFR (ml/min)<br>> 90 |
| --- | --- | --- | --- | --- |
| without kidney disease |  |  |  |  |
| 1st week | 111 | 5.23 ± 1.72 | 78.26 ± 25.14 | 129.81 ± 10.33 |
| 2st week | 108 | 5.58 ± 2.44 | 75.31 ± 23.52 | 126.37 ± 9.72 |
| 3st week | 105 | 5.04 ± 1.96 | 77.04 ± 22.27 | 128.53 ± 9.29 |
| 4st week | 104 | 5.19 ± 2.07 | 72.95 ± 24.83 | 127.96 ± 9.65 |
| P value |  | 0.877 | 0.121 | 0.177 |
| Chronic renal failure |  |  |  |  |
| 1st week | 5 | 32.08 ± 8.58 | 937.61 ± 114.62 | 14.43 ± 7.34 |
| 2st week | 5 | 30.66 ± 9.64 | 955.47 ± 141.09 | 15.96 ± 8.72 |
| 3st week | 5 | 29.79 ± 10.37 | 897.53 ± 175.48 | 21.33 ± 10.09 |
| 4st week | 5 | 31.94 ± 9.18 | 914.29 ± 163.87 | 22.86 ± 9.37 |
| P value |  | 0.981 | 0.801 | 0.152 |

BUN: blood urea nitrogen, Cr: creatinine, eGFR: glomerular filtration rate. *P* values indicate differences between 4st week and 1st week. *P* <0.05 was considered statistically significant.+

### Detection results of SARS-CoV-2 RNA in urine sediment

SARS-CoV-2 RNA in urine sediments of COVID-19-confirmed 53 patients, including 5 CRF cases, enrolled in this study was examined by real-time RT-PCR. The results showed that SARS-CoV-2 RNA in urine sediments was positive in 3 patients without renal illness before (3/48), except one patient with CRF had a positive for SARS-CoV-2 ORF 1ab (1/5).

### Mortality of 116 COVID-19-confirmed patients

As of February 13, 2020, 7 (6.03%) ARDS patients transferred to ICU died of respiratory failure. All the 7 dead patients with ARDS were over 60 years old, and the maximum age was 95 years old. None of the 7 patients exhibited ARF, but all of them had other comorbidities, including 4 patients with advanced malignant tumor, 2 patients with hypertension and coronary heart disease, and one patient with diabetes and cerebral infarction. All of the 7 patients had pulmonary consolidation and hypoxemia which was difficult to correct. Even if invasive ventilation was used, they died from respiratory failure. It’s worth noting that all of 5 patients with CRF were survived, who did not develop to ARDS or CRF deterioration.

## Discussion

The first step in SARS-CoV-2 infection is to bind to the host cell receptor and enter the cells. Recent a study shows that the common ancestor of SARS-CoV-2 and SARS-CoV is similar to bat coronavirus HKU9-1 ^[13]^. These coronaviruses have a three-dimensional structure of spike protein, which is closely bound to human cell receptor angiotensin converting enzyme II (ACE2). Therefore, the cells with ACE2 expression may act as target cells and be susceptible to COVID-19 infection, such as type II alveolar cells (AT2) in the lung ^[14]^. It should be noted that ACE2 protein has been proved to have an abundant expression in many kinds of cells, such as intestinal epithelial cells, renal tubular epithelial cells, alveolar epithelial cells, heart, artery smooth muscle cells and gastrointestinal system ^[15]^. Therefore, it is reasonable to speculate that SARS-CoV-2 may invade the lung, upper respiratory tract, ileum, heart and kidney, which may lead to dyspnea, diarrhea, acute heart injury and acute renal failure, especially in the case of viremia.

Recently, a study on kidney functions in 59 patients infected by SARS-CoV-2 was reported ^[16]^. It was found that 63% (32/51) of the patients exhibited proteinuria, 19% (11/59) and 27% (16/59) of the patients had an elevated level of plasma creatinine and urea nitrogen respectively. Moreover, the CT scan showed radiographic abnormalities of the kidneys in 100% (27/27) of the patients. Therefore, it was concluded that renal impairment is common in COVID-19 patients, which may be one of the major causes of the illness by the virus infection and also may contribute to multiorgan failure and death eventually.

In this study, the effects of SARS-CoV-2 infection on renal function were explored through analyzing the clinical data of 116 hospitalized COVID-19-confirmed patients. However, the results of renal impairment in COVID-19 patients were not observed in this study. Although 12 patients (10.8%) without kidney disease showed mild elevation of urea nitrogen or creatinine, and 8 patients (7.2%) showed trace or 1+ albuminuria after infection with the virus and during the treatment of pneumonia. These 111 COVID-19-confirmed patients without basic kidney disease did not develop obvious abnormal renal function during the treatment of pneumonia. It was found that SARS-CoV-2 infection does not cause significant renal damage. In addition, they gradually returned to normal after a follow-up and did not receive special treatment for the kidneys. The abnormal renal function is supposed as secondary injury duo to hypoxemia in these patients.

A previous study, Wang et al. reported the clinical characteristics of 138 hospitalized COVID-19-confirmed cases in a study. The data showed that the value of both blood urea nitrogen [4.4 (3.4-5.8) mmol/L (Median (IQR))] and creatinine [72 (60-87) µmol/L (Median (IQR))] were within the normal range ^[10]^. Guan et al. presented also the data of clinical characteristics of 1,099 patients confirmed with COVID-19 from 552 hospitals in 31 provinces/provincial municipalities in a study. From this study, the renal function showed that the patients’ number of Creatinine ≥ 133 µmol/L were 12/752 (1.6%) ^[18]^. Data from above two studies suggested that acute renal impairment was uncommon in COVID-19, and SARS-CoV-2 infection does not significantly cause obvious azotemia and acute renal injury.

Based on the clinical, pathologic study, and laboratory features of SARS-CoV infection in SARS patients in 2003, the data showed that acute renal impairment was uncommon, but carried a formidably high mortality (91.7%, 33 of 36 cases) ^[17]^. In this study, all of the patients without basic kidney disease showed no obvious abnormality of renal function during the hospitalization of COVID-19, and none of the patients exhibited ARF. Be cause high homology of SARS-CoV-2 and SARS-CoV. The results of this study were similar and consistent with the presentation of renal function injury in SARS. In this study, we also observed that the patients with chronic renal failure (CRF) who were undergone regular continuous renal replacement therapy (CRRT) were infected with SARS-CoV-2, and confirmed as COVID-19. Except regular CRRT, the monitoring of renal function indicators showed stable, without exacerbation of CRF during the course of treatment of COVID-19. The re-examination of CT showed that pulmonary inflammation was gradually absorbed. Unlike a formidably high mortality in SARS complicated with renal impairment, none of the patients died from the aggravation of CRF or from COVID-19 itself caused by infection with SARS-CoV-2. It was also suggested that CRRT plays an important role in the treatment of COVID-19 complicated with CRF.

CRRT is a mode of renal replacement therapy for hemodynamically unstable, fluid overloaded patients and the patients with sepsis and septic shock in management of ARF, especially in the ICU setting ^[19]^. CRRT initiated for ARF in critically ill patients should serve as a renal ‘replacement’ therapy mimicking as artificial kidney support. It should enhance recovery of the native kidneys with prevention of hyperkalemia, hyper/ hyponatremia, acidosis/alkalosis and rapid correction of pulmonary/peripheral edema by gradual and consistent removal of extra fluid retained in the body. CRRT also can play a role in removing inflammatory mediators and improving immune function in critical patients ^[20]^. Nevertheless, the renal function of patients with COVID-19 needs to be monitored regularly, especially in patients with elevated plasma creatinine. In the event of signs of ARF, potential interventions, including CRRT, should be used to protect renal function as early as possible.

## Data Availability

The data used to support the findings of this study are available from the corresponding author upon request.

